# *Streptococcus pneumoniae* re-emerges as a cause of community-acquired pneumonia, including frequent co-infection with SARS-CoV-2, in Germany, 2021

**DOI:** 10.1101/2022.12.15.22282988

**Authors:** Juliane Ankert, Stefan Hagel, Claudia Schwarz, Kaijie Pan, Liz Wang, Christof von Eiff, Bradford D. Gessner, Christian Theilacker, Mathias W. Pletz

## Abstract

**Background:** The COVID-19 pandemic and the associated containment measures had a substantial impact on pathogens causing pneumonia in adults. The objective of this study was to determine the etiology of hospitalized community-acquired pneumonia (CAP) among adults in Germany in 2021, the second year of the COVID-19 pandemic.

**Methods:** Since January 2021, this on-going, prospective, population-based surveillances study enrolled adult patients with clinically and radiographically confirmed CAP at three hospitals in Thuringia, Germany, serving a population of approximately 280,000. Urine samples were collected from patients and tested for *S. pneumoniae* using the pneumococcal urinary antigen test (PUAT, BinaxNOW *S. pneumoniae*) and the proprietary serotype-specific urinary antigen detection (UAD) assays. Nasopharyngeal swabs were tested for 10 respiratory viruses by PCR.

**Results:** A total of 797 patients were enrolled, of whom 760 were included in the analysis. The median age of patients with CAP was 67 years; in-hospital case-fatality rate was 8.4%. A respiratory pathogen was detected in 553 (72.8%) patients. The most common pathogen was SARS-CoV-2 (n=498, 68.2%), followed by *S. pneumoniae* (n=40, 6.4%). Serotypes contained in the 13-valent, 15-valent and 20-valent pneumococcal conjugate vaccine were detected in 42.5%, 45.0%, and 70.0% of the pneumococcal CAP cases. Between the first and second half of 2021, the proportion of CAP cases associated with *S. pneumoniae* increased from 1.1% to 5.6% in patients aged 18-59 years and from 2.5% to 12.4% in those aged ≥60 years; coinfection of SARS-CoV-2 and *S. pneumoniae* among COVID-19 patients increased from 0.7% (2/283 cases) to 6.0% (13/215) in patients aged ≥18 years, and from 1.0% (2/195) to 8.7% (11/127) in those aged ≥60 years.

**Conclusion:** In Germany, the proportion of CAP cases associated with *S. pneumoniae* rebounded to a near-pandemic level in the second half of 2021 and many pneumococcal infections occurred in patients with COVID-19. Vaccination uptake against respiratory pathogens, including *S. pneumoniae*, should be strengthened.

---

Following the emergence of the COVID-19 pandemic in early 2020, many countries implemented containment measures to curb the spread of SARS-CoV-2. These containment measures were associated with a substantial reduction in the activity of other respiratory viruses as well as bacterial disease such as pneumococci [1, 2]. Consequently, as containment measures were gradually withdrawn, a rebound of RSV disease in children [3] and increases in pneumococcal diseases (IPD) were observed [4]. However, little is known how increasing population-immunity against SARS-CoV-2, the circulation of new SARS-CoV-2 variants, and withdrawal of containment measures have impacted the microbiology of community-acquired pneumonia (CAP) over time. Here, we describe the etiology of hospitalized CAP among adults in Germany during 2021, the second year of the COVID-19 pandemic. In 2021, Germany had implemented strict COVID-19 containment measures, which were gradually withdrawn over the year and re-implemented before 2022 [5].

This ongoing prospective, population-based surveillance study is conducted since January 4, 2021, at one community and two large tertiary care hospitals in Thuringia, Germany, serving a population of about 280,000 inhabitants. Adult patients with suspicion of lower respiratory tract infection admitted to study hospitals were screened for eligibility and individuals with documented or clinically suspected pneumonia were enrolled. Patients included in this analysis had (1) radiologically-confirmed CAP diagnosed within 48 hours of hospital admission, (2) per-protocol nasopharyngeal swabs (NPS) and urine samples collected, and (3) information on hospital discharge disposition available. Study staff collected data on patient characteristics, medical history, hospital course, and standard of care microbiologic testing. NPS were tested by PCR for 10 respiratory viruses (SARS-CoV-2, respiratory syncytial virus [RSV], influenza virus, parainfluenza virus, human metapneumovirus, rhinovirus, human endemic coronavirus, adenovirus, enterovirus, bocavirus). Urine samples were tested with the pneumococcal urinary antigen test (PUAT, BinaxNOW *S. pneumoniae*) and with proprietary serotype-specific urinary antigen detection (UAD) assays [6, 7]. UAD assays detect all serotypes included in currently licensed vaccines. All data were descriptively summarized.

A total of 2084 patients were screened, 797 enrolled, and 760 included in this analysis. The median age of patients with radiologically-confirmed CAP was 67 years (Q1, Q3: 58, 79) and 57.6% were male. Approximately three quarters (75.9%) of patients had at least one comorbidity. The uptake for same season influenza vaccine, cumulative uptake of 23-valent pneumococcal polysaccharide vaccine (PPV23), and the 13-valent pneumococcal conjugate vaccine (PCV13) were 23.4%, 25.0% and 3.0%, respectively. The uptake for ≥1 dose of a COVID-19 vaccine was 14.3% in the first half of 2021 and increased to 50.6% in the second half. The median hospital length of stay was 8 days, 122 patients (16.1%) were admitted to the ICU, 43.7% had a pneumonia severity index (PSI) grade of IV or V, and in-hospital case-fatality rate was 8.4%.

A respiratory pathogen was detected in 553 (72.8%) patients with CAP. The most frequent pathogen was SARS-CoV-2 (n=498, 68.2%), followed by *S. pneumoniae* (n=40, 6.4%), rhinovirus and parainfluenza virus (n=16, 2.3% each), and RSV (n=14, 2.0%) (**Figure 1C**). Other viruses were infrequent (≤1.0% each), and no case of influenza was identified. The percentage of CAP patients with SARS-CoV-2 detection was higher in patients aged 18 to 59 years (n=176, 79.3%) than those aged ≥60 years (n=322, 63.4%). Conversely, *S. pneumoniae* was more frequently detected in patients aged ≥60 years (n=33, 7.8%) than in patients aged 18 to 59 years (n=7, 3.6%). Of the 40 pneumococcal CAP cases, PCV13-, PCV15-, PCV20- and PPV23-included serotypes, accounted for 42.5% (n=17), 45.0% (n=18), 70.0% (n=28), and 73% (n=29). The most frequent serotypes were 3 (n=5), 8 (n=4), and 19A/6A/11A (3 detections each).

**Figure 1.**
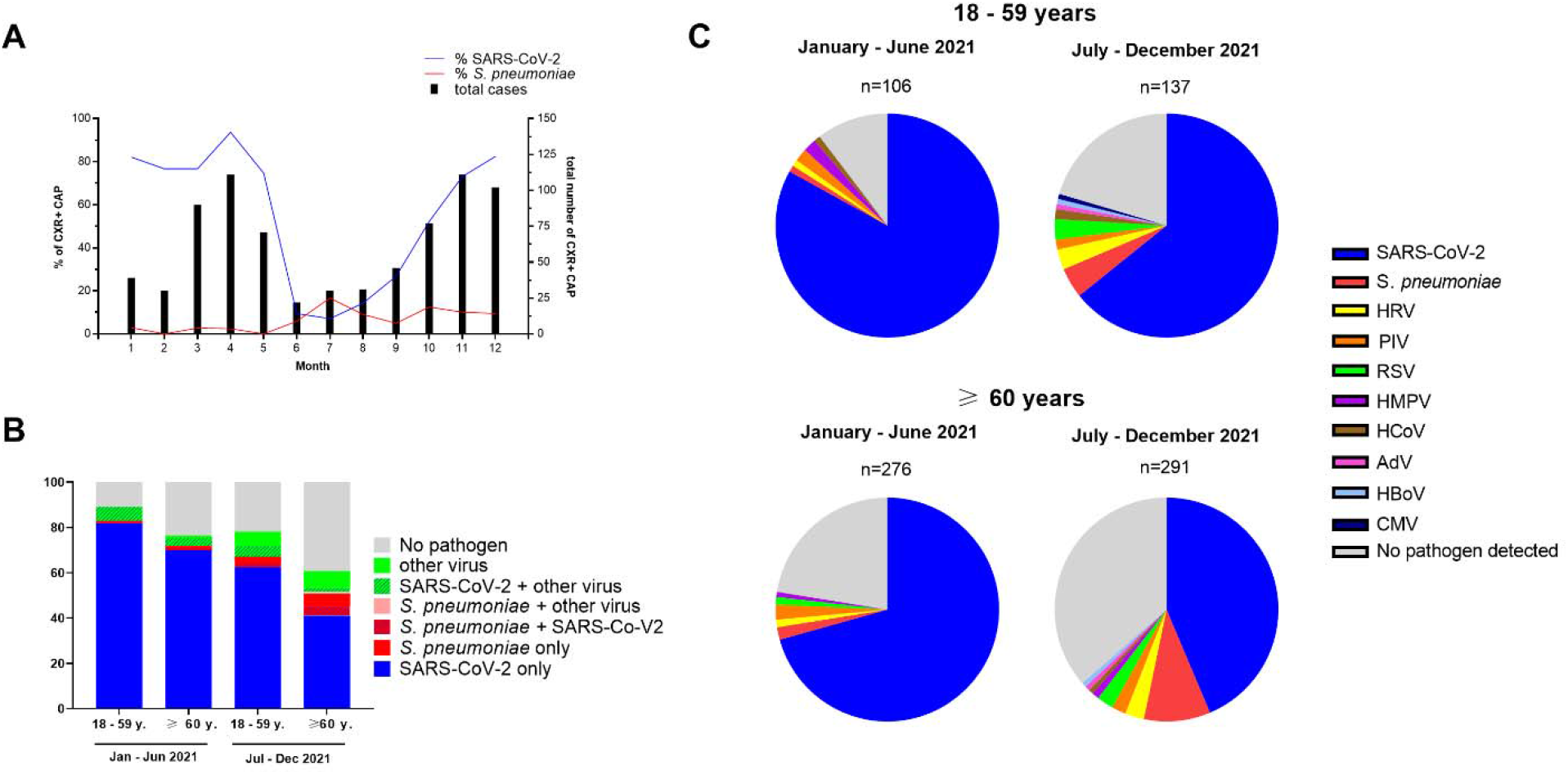
Pathogen detection among adults with community-acquired pneumonia requiring hospitalization in Thuringia, Germany in 2021. For all patients enrolled in the study, a nasopharyngeal swab and urine was collected and multiplex PCR for 10 respiratory viruses and urinary antigen tests for the C-wall polysaccharide (BinaxNow *S. pneumoniae*) and serotype antigens (urinary antigen detection assay) were performed. Results from the per-protocol and standard of care testing are shown. **A** -Total number of enrolled adults with radiologically-confirmed, hospitalized CAP, and proportion of patients with detection of SARS-CoV-2 and *S. pneumoniae* between January 4, 2021, and December 31, 2021, by month. **B** - Proportion of CAP due to SARS-CoV-2, *S. pneumoniae* or other respiratory viruses, by age group and study period. **C** - Proportion pathogens detected, according to age group and study period. n indicates the total number of pathogen detections. Abbreviations: SARS-CoV-2 - severe acute respiratory syndrome coronavirus 2, *S. pneumoniae – Streptococcus pneumoniae*, HRV - human rhinovirus, PIV - parainfluenza virus 1 - 4, HMPV - human metapneumovirus, HCoV - human coronavirus (229E, OC43, NL63), RSV - respiratory syncytial virus A and B, AdV - adenovirus, HBoV - human bocavirus, CMV - cytomegalovirus.

The etiology of CAP changed substantially throughout 2021 (**Figure 1**). The monthly proportion of CAP cases associated with SARS-CoV-2 ranged from 76.7% to 93.7% between January and May, dipped to 7.1% in July, and then rose continuously to 82.4% until December (**Figure 1A**). *S. pneumoniae* caused 0.0% to 2.9% of CAP between January and May and accounted for 5.9% to 16.7% afterwards (**Figure 1A**). The proportion of CAP cases associated with SARS-CoV-2 decreased from the first to second half of 2021 in both older adults (from 74.7% to 51.4%) and younger adults (from 88.0% to 72.1%) (**Figure 1B**). By contrast, the proportion of CAP cases associated with *S. pneumoniae* increased between the first and second half of 2021 in older adults from 2.5% to 12.4% and in younger adults from 1.1% to 5.6% (**Figure 1B**). Similarly, respiratory viruses other than SARS-CoV-2 increased between the first and second half of 2021 from 5.3% to 11.3%. Coinfection of SARS-CoV-2 and *S. pneumoniae* occurred in 2/283 (0.7%) of patients in the first half of 2021, but in 13/215 (6.0%) in adults ≥18 years and to 11/127 cases (8.7%) in adults ≥60 years in the second half of 2021 (**Figure 1B**). Thirteen out of a total of 34 (38.5%) pneumococcal detections in the second half of 2021 were in patients with COVID-19 (**Figure 1B**). Coinfections of SARS-CoV-2 with other respiratory viruses remained unchanged at around 5% over the year 2021.

In this pneumonia surveillance study, we describe changes with CAP etiology in Germany during 2021, covering the time of the spread of the COVID-19 Wuhan strain, and the subsequent waves of Alpha and Delta variants [8]. SARS-CoV-2 was the predominant cause of CAP in 2021, and the percentage of CAP due to *S. pneumoniae* rose from very low levels in early 2021 to near pre-pandemic levels among older patients in the second half of 2021 [9], with 12.4% of CAP cases caused by *S. pneumoniae* and 78.5% of these due to PCV20 serotypes. Previous studies reported that *S. pneumoniae* coinfection was rare among COVID-19 patients but if present, associated with high case-fatality [10, 11]. In the second half of 2021, *S. pneumoniae* was detected in 8.7% of older adults with COVID-19, a coinfection rate higher than recently reported for influenza [12]. The re-emergence of *S. pneumoniae* may be related to several factors, including changes in social mixing with relaxation of containment measures and healthcare seeking behavior, and differences in the interference of individual SARS-CoV-2 variants with other respiratory pathogens [13]. Underestimation of pneumococcal CAP by previous studies may also be related to the low sensitivity of the standard-of-care culture methods compared to urinary antigen testing used in this study.

The main strength of this study lies in the high testing rates of > 80% for 11 respiratory pathogens. A limitation of the study is that of 2084 screened patients, only 797 (38%) were enrolled. The most common reason for non-enrollment was a lack of capacity to provide consent, reducing the generalizability of our results to severely ill, debilitated patients. Also, the roll-out of the COVID-19 vaccination campaign in 2021 preferentially targeted older adults and this may bias comparisons across age groups.

*S. pneumoniae*, the leading bacterial cause of CAP prior to the COVID-19 pandemic, was reduced during the early months of 2021 but rebounded to near pre-pandemic levels after the summer of 2021, affecting both COVID-19 and non-COVID-19 CAP patients in Germany. This rebound occurred in the absence of influenza circulation. With a more diverse spectrum of CAP etiologies emerging, it will be critical to strengthen vaccination uptake not only for SARS-CoV2, but also for the currently re-emerging influenza [14] and *S. pneumoniae* [15]. In addition, clinical treatment guidelines for COVID-19 patients should consider inclusion of empiric treatment for *S. pneumoniae*, a practice that is currently discouraged based on low pneumococcal coinfection rates reported during the earlier phase of the COVID-19 pandemic [16].

## Data Availability

All data produced in the present study are available upon reasonable request to the authors.

## Acknowledgements

We would like to thank the research team at the University Medical Center Jena, including Sebastian Weis, Christina Bahrs, Anne Moeser, Bettina Löffler, and Steffi Kolanos, the research team at the SRH Klinik Gera, including Dagmar Täuscher, Nancy Schmidt, Nicole Haupt, Sabine Sell, Anne Meinzenbach, Melissa Rohde, Romy Anger, Mandy Michaelis, Mandy Wießner, Kristin Zoeger, and Julia Schulze, and the research team at the SRH Klinik Suhl, including Mohamed Ahmed El Sebai, Christian-Marian Andrei, Anastasia Mihali, Mara-Viviana Crasnic, Nicole Gerlach, Gabriela Günther, Nadine Marr, and Stefanie Resiger for their support with the study. Medical writing support was provided by Qi Yan (Pfizer Inc.).

## References

1. Olsen SJ, Winn AK, Budd AP, Prill MM, Steel J, Midgley CM, Kniss K, Burns E, Rowe T, Foust A, Jasso G, Merced-Morales A, Davis CT, Jang Y, Jones J, Daly P, Gubareva L, Barnes J, Kondor R, Sessions W, Smith C, Wentworth DE, Garg S, Havers FP, Fry AM, Hall AJ, Brammer L, Silk BJ. Changes in Influenza and Other Respiratory Virus Activity During the COVID-19 Pandemic - United States, 2020-2021. MMWR Morb Mortal Wkly Rep 2021: 70(29): 1013–1019.

2. Brueggemann AB, Jansen van Rensburg MJ, Shaw D, McCarthy ND, Jolley KA, Maiden MCJ, van der Linden MPG, Amin-Chowdhury Z, Bennett DE, Borrow R, Brandileone M-CC, Broughton K, Campbell R, Cao B, Casanova C, Choi EH, Chu YW, Clark SA, Claus H, Coelho J, Corcoran M, Cottrell S, Cunney RJ, Dalby T, Davies H, de Gouveia L, Deghmane A-E, Demczuk W, Desmet S, Drew RJ, du Plessis M, Erlendsdottir H, Fry NK, Fuursted K, Gray SJ, Henriques-Normark B, Hale T, Hilty M, Hoffmann S, Humphreys H, Ip M, Jacobsson S, Johnston J, Kozakova J, Kristinsson KG, Krizova P, Kuch A, Ladhani SN, Lâm T-T, Lebedova V, Lindholm L, Litt DJ, Martin I, Martiny D, Mattheus W, McElligott M, Meehan M, Meiring S, Mölling P, Morfeldt E, Morgan J, Mulhall RM, Muñoz-Almagro C, Murdoch DR, Murphy J, Musilek M, Mzabi A, Perez-Argüello A, Perrin M, Perry M, Redin A, Roberts R, Roberts M, Rokney A, Ron M, Scott KJ, Sheppard CL, Siira L, Skoczyńska A, Sloan M, Slotved H-C, Smith AJ, Song JY, Taha M-K, Toropainen M, Tsang D, Vainio A, van Sorge NM, Varon E, Vlach J, Vogel U, Vohrnova S, von Gottberg A, Zanella RC, Zhou F. Changes in the incidence of invasive disease due to Streptococcus pneumoniae, Haemophilus influenzae, and Neisseria meningitidis during the COVID-19 pandemic in 26 countries and territories in the Invasive Respiratory Infection Surveillance Initiative: a prospective analysis of surveillance data. The Lancet Digital Health 2021: 3(6): e360–e370.

3. Danino D, Ben-Shimol S, van der Beek BA, Givon-Lavi N, Avni YS, Greenberg D, Weinberger DM, Dagan R. Decline in Pneumococcal Disease in Young Children During the Coronavirus Disease 2019 (COVID-19) Pandemic in Israel Associated With Suppression of Seasonal Respiratory Viruses, Despite Persistent Pneumococcal Carriage: A Prospective Cohort Study. Clin Infect Dis 2022: 75(1): e1154–e1164.

4. Bertran M, Amin-Chowdhury Z, Sheppard CL, Eletu S, Zamarreño DV, Ramsay ME, Litt D, Fry NK, Ladhani SN. Increased Incidence of Invasive Pneumococcal Disease among Children after COVID-19 Pandemic, England. Emerg Infect Dis 2022: 28(8): 1669–1672.

5. Our World in Data. Oxford Coronavirus Government Response Tracker (OxCGRT) [cited 27. Oct 2022]; Available from: https://ourworldindata.org/covid-stringency-index

6. Pride MW, Huijts SM, Wu K, Souza V, Passador S, Tinder C, Song E, Elfassy A, McNeil L, Menton R, French R, Callahan J, Webber C, Gruber WC, Bonten MJ, Jansen KU. Validation of an immunodiagnostic assay for detection of 13 Streptococcus pneumoniae serotype-specific polysaccharides in human urine. Clin Vaccine Immunol 2012: 19(8): 1131–1141.

7. Kalina WV, Souza V, Wu K, Giardina P, McKeen A, Jiang Q, Tan C, French R, Ren Y, Belanger K, McElhiney S, Unnithan M, Cheng H, Mininni T, Giordano-Schmidt D, Gessner BD, Jansen KU, Pride MW. Qualification and Clinical Validation of an Immunodiagnostic Assay for Detecting 11 Additional Streptococcus pneumoniae Serotype-specific Polysaccharides in Human Urine. Clin Infect Dis 2020: 71(9): e430–e438.

8. Institute for Infectious Diseases and Infection Control University Hospital Jena G. Monthly SARS-CoV-2 lineages (Thuringia). [cited 27. Oct. 2022]; Available from: https://charts.mongodb.com/charts-routine-sequencing-sars-c-amykg/public/dashboards/e9453286-1dce-4202-9423-a8459e3962f8

9. Bahrs C, Kesselmeier M, Kolditz M, Ewig S, Rohde G, Barten-Neiner G, Rupp J, Witzenrath M, Welte T, Pletz MW, Group CS. A longitudinal analysis of pneumococcal vaccine serotypes in pneumonia patients in Germany. Eur Respir J 2022: 59(2).

10. Langford BJ, So M, Raybardhan S, Leung V, Westwood D, MacFadden DR, Soucy JR, Daneman N. Bacterial co-infection and secondary infection in patients with COVID-19: a living rapid review and meta-analysis. Clin Microbiol Infect 2020.

11. Amin-Chowdhury Z, Aiano F, Mensah A, Sheppard C, Litt D, Fry NK, Andrews N, Ramsay ME, Ladhani SN. Impact of the COVID-19 Pandemic on Invasive Pneumococcal Disease and Risk of Pneumococcal Coinfection with SARS-CoV-2: prospective national cohort study, England. Clin Infect Dis 2020.

12. Bartley PS, Deshpande A, Yu PC, Klompas M, Haessler SD, Imrey PB, Zilberberg MD, Rothberg MB. Bacterial coinfection in influenza pneumonia: Rates, pathogens, and outcomes. Infection control and hospital epidemiology 2022: 43(2): 212–217.

13. Hyams C, Begier E, Garcia Gonzalez M, Southern J, Campling J, Gray S, Oliver J, Gessner BD, Finn A. Incidence of acute lower respiratory tract disease hospitalisations, including pneumonia, among adults in Bristol, UK, 2019, estimated using both a prospective and retrospective methodology. BMJ Open 2022: 12(6): e057464.

14. Australian Government Department of Health ang Aged Care. Australian Influenza Surveillance Report - No. 14, 2022, reporting fortnight: 26 September to 09 October 202. 2022 [cited 27. Oct 2022]; Available from: https://www1.health.gov.au/internet/main/publishing.nsf/Content/cda-surveil-ozflu-flucurr.htm/$File/flu-14-2022.pdf

15. The Academy of Medical Sciences. COVID-19: Preparing for the future. Looking ahead to winter 2021/22 and beyond. 2021 [cited August 10, 2021]; Available from: https://acmedsci.ac.uk/filedownload/4747802

16. National Institutes of Health. COVID-19 Treatment Guidelines Panel. Coronavirus Disease 2019 (COVID-19) Treatment Guidelines. [cited 20. October 2022]; Available from: at https://www.covid19treatmentguidelines.nih.gov/.

